# Postpartum and Youth Depression in the Context of Vitamin D Supplementation: Systematic Review and Meta-analysis

**DOI:** 10.1101/2025.08.14.25333707

**Authors:** Joseph P. Nano, William A. Catterall, Salar Khaleghzadegan, Camilo A. Castelblanco, Heather B. Blunt, Renata W. Yen

## Abstract

**Background:** We conducted a systematic review and meta-analysis to assess the effect of vitamin D supplementation on depression among adolescents and young adults compared with placebo and baseline.

**Methods:** We searched databases and reference lists from inception to May 2024 for English-language studies, including cohort studies, case studies, and randomized clinical trials. Study quality was assessed using the Cochrane RoB2 tool and the Newcastle-Ottawa Scale. Random-effects meta-analyses estimated the standardized mean difference (SMD) in depression scores (primary outcome) and anxiety scores (secondary outcome). Heterogeneity was assessed using the I² statistic.

**Results:** Fifteen studies (2010–2024) from 7,638 citations met inclusion criteria. The mean sample size was 5,271 (range: 38–74,840). Meta-analysis of nine studies showed that vitamin D supplementation significantly reduced depression scores versus placebo (SMD: −0.43; 95%CI: −0.75 to −0.12; p=0.007; I²=78%). Of six studies not included in the meta-analysis, five reported significant associations between supplementation and lower depression. Subgroup effects were observed for postpartum women (SMD: −0.55; 95%CI: −1.04 to −0.06; p<0.05; I²=83%) and young adults (SMD: −1.38; 95%CI: −1.65 to −1.10; p<0.001; I²=76%). Meta-analysis of four studies found no significant association with anxiety (SMD: −0.31; 95%CI: −0.75 to 0.12; p=0.16; I²=77%).

**Limitations:** Variability in study sample sizes may affect interpretation and generalizability.

**Conclusions:** Vitamin D supplementation was associated with lower depression scores, with effects varying by subgroup.

## INTRODUCTION

Depression is a major public health concern, affecting approximately 5% of adults worldwide (about 280 million people).^1^ It can lead to severe mood disorders that disrupt appetite, energy, and thoughts.^1^ There are gender disparities in depression that start during adolescence and may continue throughout adulthood.^2,3^ While depression manifests in various forms, including major depressive disorder, bipolar disorder, seasonal affective disorder, and peripartum (postpartum) depression,^4–6^ In March 2020, the COVID-19 pandemic exacerbated the burden of depression and mental health illness among adolescents and adults worldwide.^7–9^ Vitamin D, also known as cholecalciferol (vitamin D_3_) and ergocalciferol (vitamin D_2_), plays a critical role in regulating calcium metabolism to protect against the development of various diseases.^10,11^ Vitamin D supplements are primarily available in the form of vitamin D_3_, which is the more effective form at raising and maintaining vitamin D levels in the body.^12^ Vitamin D is a lipid-soluble compound with receptors (VDR) distributed across the brain parenchyma, including neurons in the cerebral cortex and hypothalamus.^13,14^ A key cellular role of vitamin D is the regulation of calcium levels in the brain, which is critical for human development.^15,16^ Several postulates, including the phenotypic stability hypothesis, suggest that decreased vitamin D levels are one factor that enables abnormal increases in Ca^2+^ concentrations associated with depression.^17^ Additionally, vitamin D functions as a neuroactive steroid that plays a role in neuroimmunomodulation and production of antioxidants, which makes it biologically plausible to be associated with depression.^18,19^

The relationship between vitamin D levels and depression remains controversial, with some studies reporting mixed results regarding their association.^20–24^ In addition, studies that investigated the association between vitamin D serum levels and postpartum depression found mixed results.^25–28^ Other studies that investigated the association between vitamin D supplements and depression also revealed mixed results.^29–32^ Furthermore, a recent review that investigated the neural activities of vitamin D and its association with depression found that lower vitamin D serum levels are associated with an increased risk for depression.^33^ However, there remains a gap in literature investigating the association between vitamin D supplementation and depression among young adults and depression, including postpartum depression. Also, a recent systematic review and meta-analysis study that focused on the association between vitamin D serum levels and depression among adolescents and young adults revealed that more studies are showing an inverse correlation between vitamin D serum levels and depression.^34^

To address these gaps, we aimed to conduct a systematic review and meta-analysis to investigate whether vitamin D supplementation intake, compared to placebo, is associated with decreased depression scores among adolescents and young adults, including postpartum women.

## METHODS

### Eligibility criteria

#### Study design

We created a protocol for this systematic review and meta-analysis in July 2024 to guide the inclusion of studies. We registered our protocol in August 2024 on Open Science Framework (see **Appendix 7** for modifications made since the review was registered). We followed standard analysis procedures outlined by the Cochrane Handbook and the PRISMA guidelines when reporting methods and results.^35,36^

#### Inclusion criteria

Before analyzing the selected studies, we established inclusion criteria based on study design, population of interest, exposure, control, and outcome (see **Appendix 1** for specific inclusion criteria and outcomes of interest). Our population of interest was adolescents (aged 10-17 years)^37^, and young adults (aged 18-40 years)^38,39^. Since pregnancy typically occurs during young adulthood (women aged 18-40 years)^40^ and postpartum depression represents a distinct type of depressive episode influenced by unique biological and social factors, we included studies that investigated the effect of vitamin D supplementation on postpartum depression and examined how outcomes in this subgroup differed from other young adult subgroups. Within our young adult population, we categorized participants as maternal (postpartum women) and non-maternal (all other young adult subgroups). We included randomized controlled trials (RCT), cohort studies, and case studies in any country (see Appendix 2 for PubMed search terms used) (see **Appendix 3** for search strategies). Included studies had to have vitamin D supplementation as an intervention tool and placebo as a comparison group for randomized controlled trials or baseline scores within each group for cohort and case studies. We limited our search to studies in English to avoid potential translation bias. We excluded studies that did not match our language or age criteria (**Appendix 1**).

### Outcomes

Our primary outcome was depression, assessed using validated depression measures. We selected secondary outcomes related to the benefits and harms of vitamin D supplementation: weight, body mass index (BMI), alcohol consumption, anxiety, physical activity status, and smoking status. However, we found sufficient data only for anxiety as a secondary outcome. These studies used validated measures to assess anxiety.

### Data sources

In collaboration with a biomedical reference librarian (HBB), we (JN and WC) developed a list of keywords and subject headings in MEDLINE (via PubMed) (see **Appendix 3** for search strategies). We conducted a preliminary search in MEDLINE (via PubMed) to explore whether a sufficient number of studies existed to address our research question. Our search included all dates up to the search date, May 29, 2024. We used exploded MeSH terms and keywords to generate search results regarding the association between vitamin D supplementation and depression. We used the Boolean “AND” to find the intersection between themes. We performed full electronic searches in MEDLINE (via PubMed), The Cochrane Library, Cumulative Index to Nursing and Allied Health Literature (CINAHL), PsycINFO, and ClinicalTrials.gov.^41–44^

### Selection process

Two independent reviewers (JN and WC) used Rayyan to screen studies in two stages. First, they reviewed titles and abstracts to exclude studies that did not meet the inclusion criteria. Second, included studies were retrieved in full text and assessed in detail against the inclusion criteria. We resolved data discrepancies after review through discussion.

### Data collection process

Two independent reviewers (JN and WC) used a piloted standardized data collection form and performed an independent double data extraction (see **Appendix 4** for blank data collection form). Reviewers extracted information from the included studies concerning the outcome, authors, publication year, country, type of study design, aim(s) and research questions, sample size, follow-up, and control condition.

### Synthesis methods

We performed a meta-analysis in RevMan (Review Manager version 5.4.1), using a random-effect model. We examined the association between vitamin D supplementation and depression. We assessed heterogeneity using the χ2 test and I^2^ test.^45^ The Cochrane Handbook’s guidelines were used to interpret heterogeneity as follows: I^2^ of 0–40% indicated potentially not important heterogeneity, 30–60% indicated moderate heterogeneity, 50–90% indicated potentially considerable heterogeneity, and 75–100% indicated considerable heterogeneity.^46^ We considered a threshold of I^2^ > 50% to indicate significant heterogeneity. A p-value threshold of <0.1 was used to determine statistical significance. For our continuous outcomes of depression and anxiety, we calculated the pooled standardized mean difference. We conducted two types of meta-analysis based on the data available: **(1)** between-group meta-analysis and **(2)** within-group analysis. First, our primary method for answering our research question was to conduct a between-group meta-analysis. For papers that did not include a mean difference for either the primary or secondary outcome of interest, we calculated these values using the data available in each respective study.^47–53^ We computed the mean change in depression scores from baseline to 5-month follow-up between vitamin D and placebo groups (e.g., mean difference in change scores = [Follow-up mean score - Baseline mean score]_Vitamin D_ - [Follow-up mean score-Baseline mean score]_Placebo_; negative change score reflects a reduction in depression scores) (See **Appendix 6A** for mathematical equations used for this type of analysis). Also, we calculated the standard deviation for the change scores based on the recommendations of Chi and colleagues and used a mathematical equation provided by Thomson and colleagues (**Appendix 6B**).^54–56^ For the SD equation, we assumed correlation coefficient to equal 0.5 due to the insufficient published data available and conducted sensitivity analysis using different values (e.g., 0.3, 0.5, 0.7) to check how the results change. A value of 0.5 is commonly used in meta-analyses as a conservative midpoint when the true correlation is unknown or when imputing standard deviations from other studies is unavailable.^57–60^ We used this strategy to assess whether vitamin D supplementation leads to a greater reduction in depression scores compared to placebo.

Second, for the studies that reported only “before” vs. “after” supplementation within the group that received vitamin D supplementation, we conducted a meta-analysis that focused on within-group analysis for vitamin D group baseline vs. vitamin D group follow-up to explore how depression scores change over time, specifically within the vitamin D group (e.g., baseline depression scores for vitamin D group vs. depression scores after a period of time while receiving vitamin D supplements).

For our secondary outcome of anxiety, we conducted the same between-group and within-group analysis as above.

### Assessment of Methodological Quality

Based on the study designs of the included articles, we used the Cochrane risk-of-bias tool for randomized trials version 2 (RoB 2) to assess the quality of randomized studies,^61,62^ and the Newcastle-Ottawa Quality Assessment Scale to assess the quality of non-randomized studies included in this systematic review.^61,62^ Two independent blinded reviewers (JN and WC) assessed all studies using the RoB 2 framework, which evaluates risk of bias across five domains: (1) bias arising from the randomization process, (2) bias due to deviations from intended interventions, (3) bias due to missing outcome data, (4) bias in measurement of the outcome, and (5) bias in selection of the reported result. Each domain was rated as “low risk of bias,” “some concerns,” or “high risk of bias.” Overall risk of bias judgments were generated for each study based on domain-level ratings. For non-randomized studies, two independent blinded reviewers (JN and WC) assessed studies based on three categories, receiving a certain number of stars with a maximum number of 9 stars indicating the highest quality. The three categories were selection, comparability, and outcome (for cohort studies) or exposure (for case-control studies). For example, high quality studies received 7 to 9 stars, moderate quality studies received 4 to 6 stars, and low quality studies received less than 4 stars. We resolved all disagreements after discussion.

### Reporting bias assessment

We created funnel plots using RevMan for each outcome reported by studies to perform visual examinations for heterogeneity. We visually inspected the funnel plot for asymmetry, looking specifically for a lack of symmetry around the summary effect size, which would suggest the presence of publication bias. In particular, we considered the possibility of publication bias if smaller studies with negative or nonsignificant results appeared to be missing from the lower left side of the funnel plot.

### Certainty assessment

We used the Grading of Recommendations Assessment Development and Evaluation (GRADE) approach for certainty assessment.^63^ This tool specifies four levels of certainty for a body of evidence for a given outcome: high, moderate, low, and very low. This assessment considers factors such as study design, risk of bias, consistency of results, directness of evidence, and precision of estimates. Two independent blinded reviewers (JN and WC) used GRADE assessment and resolved disagreements.

## RESULTS

### Study selection

Our initial searches included 7,638 citations (**Figure 1**). We removed duplicates and reviewed titles and abstracts, which led to 254 full-text articles. We included 15 studies in this review and 10 studies that provided sufficient continuous data for meta-analysis (**Figure 1**).

**Figure 1:**
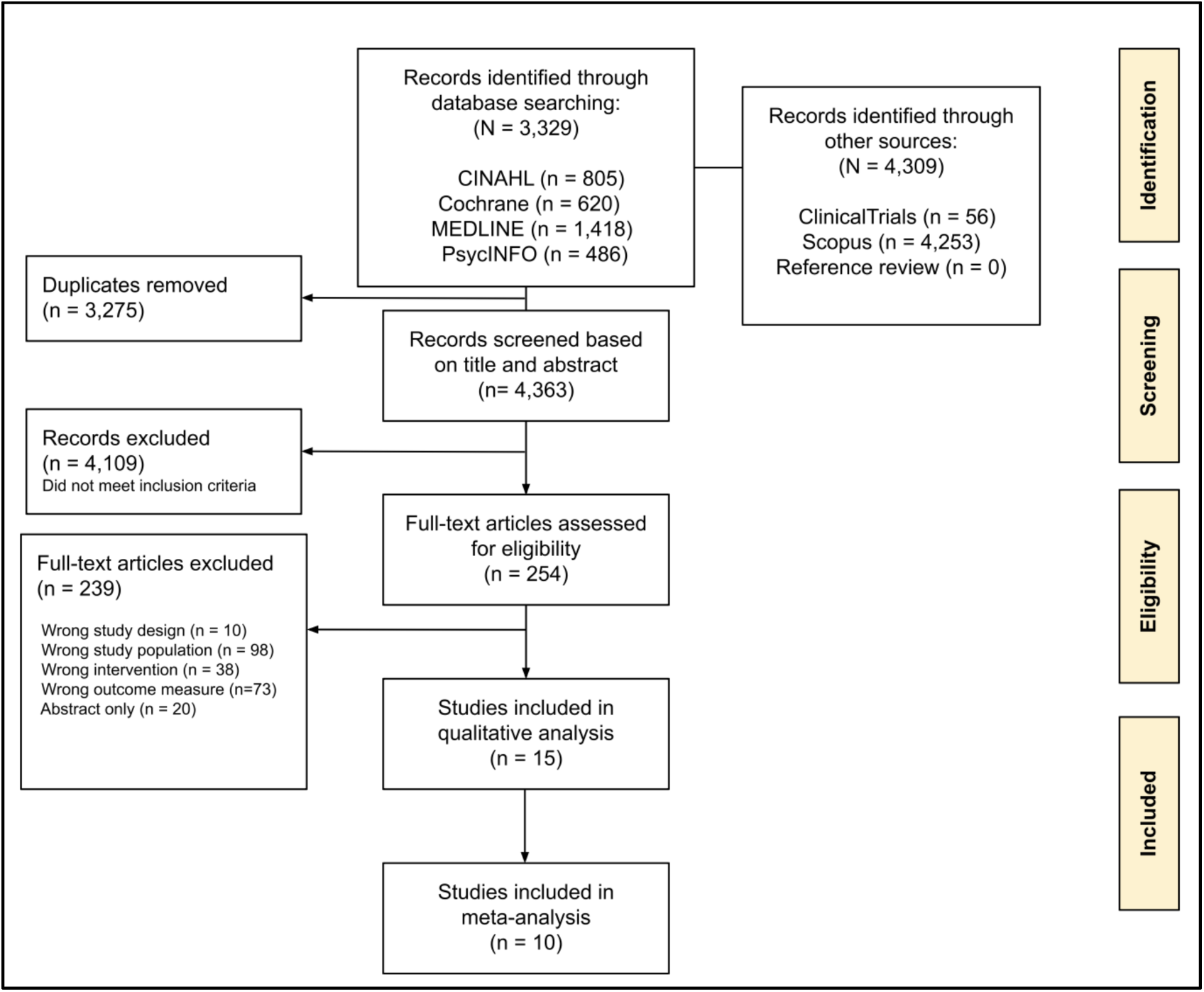
PRISMA flow chart

### Study characteristics

Our systematic review consisted of four cohort studies (26%), ten randomized clinical trial studies (67%), and one case-series study (7%). Our review included studies published between 2011 and 2024, with a mean sample size of 5,271 (range: 47 to 74,840 participations) (**Table 1**). Studies were conducted in Australia (n = 1, 7%), Germany (n = 1, 7%), India (n = 1, 7%), Iran (n = 6, 39%), Japan (n = 2, 12%), New Zealand (n = 1, 7%), Poland (n = 1, 7%), Sweden (n = 1, 7%), and the United States of America (n = 1, 7%). The population of interest in these studies were adolescents (10-18 years old) (n = 4, 27%) and young adults (18-40 years old) (n = 11, 73%), categorized as maternal during postpartum (n = 6, 55%) and all other young adult categories (e.g., non-postpartum cases) (n = 5, 45%). Two studies included reported race or ethnicity (n = 2, 12%), and the majority of studies included only women (n = 11, 73%).

**Table 1.**
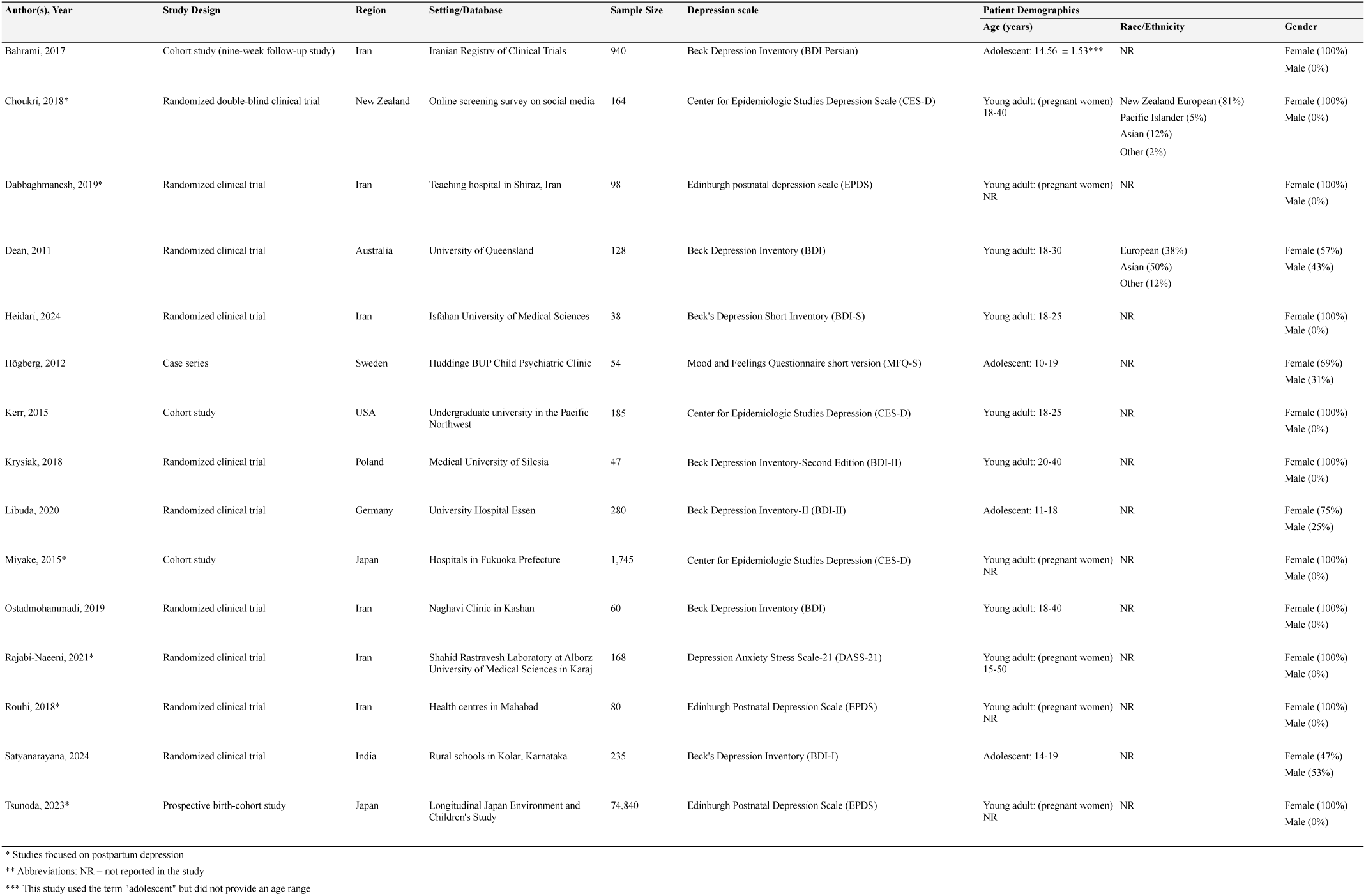
Characteristics of included studies.

### Risk of bias in studies

Version 2.0 of the Cochrane tool was used to assess the risk of bias of the eligible randomized controlled trial studies (n = 10).^64^ All trials were rated as having a low risk of bias across key domains, including randomization process, deviations from intended interventions, missing outcome data, measurement of the outcome, and selection of the reported result. Non-randomized trial studies were rated as “good” using the Newcastle-Ottawa Scale to analyze participant selection, comparability, and outcome assessment (**Appendix 10**).

### Results of individual studies

#### Validity of depression and vitamin D serum levels measurement instruments

Approximately half of the studies used either Beck Depression Inventory (BDI) (47%, n=7).^49–51,65–68^ Other studies used the Center for Epidemiologic Studies Depression Scale (CES-D) (20%, n=3),^47,69,70^ Edinburgh Postnatal Depression Scale (EPDS) (20%, n=3),^48,53,71^ Mood and Feelings Questionnaire short version (MFQ-S) (7%, n=1),^72^ and Depression Anxiety Stress Scale-21 (DASS-21) (7%, n=1).^52^

#### Association between vitamin D supplementation and depression

Eleven studies (73%) demonstrated that vitamin D supplementation decreased depression scores compared to the control group (**Table 2**). These studies reported mean sample size of 7,119 (range: 47 to 74,840) and were conducted in Japan (n=2), India (n=1), Iran (n=6), Poland (n=1), Sweden (n=1) (Table 1, Table 2). The remaining four studies found no evidence of lower depression scores as a result of vitamin D supplementation.^47,49,67,69^ These four studies were conducted in different countries (United States, Germany, Australia, New Zealand), and half of these studies used Beck Depression Inventory (BDI) (n = 2, 50%).^49,67^ The majority of these four studies were randomized clinical trials (n = 3, 75%), and their mean sample size was 189 participants (compared to mean sample size for all included studies = 5,271). The population of interest for most of these studies was young adults (n = 3,75%) (**Table 1**).^47,49,69^

**Table 2.** Association between vitamin D supplementation and depression.

| Authors, Year | Overall findings and subgroup analysis details | Results | Placebo, N. (%) | Vitamin D Supplementation, N. (%) | OR (95% CI)<sup>2</sup> | P-value<sup>1</sup> |
| --- | --- | --- | --- | --- | --- | --- |
| Vitamin D serum levels |  |  |  |  |  |  |
| Bahrami, 2017 | After nine weeks of vitamin D supplementation, there was a significant reduction in mild, moderate, and severe depression scores. However, vitamin D supplementation had no significant effect on aggression scores. These results suggest that vitamin D may improve depressive symptoms among adolescent girls, as assessed by questionnaire, but does not appear to affect aggression. | No depression, *Median (interquartile range)*<br>Mild depression<br>Moderate depression<br>Severe depression | Before vitamin D supplementation<br>5 (2-8)<br>16 (15-18)<br>23 (21-26)<br>33 (30.75-40.5) | After vitamin D supplementation<br>4 (1-9)<br>15 (9-18)<br>20 (11-24)<br>26 (9-34) | NR | 0.904<br>0.002<br>0.001<br>0.001 |
| Choukri, 2018 | No evidence of lower depression (P = 0.339), lower anxiety (P = 0.862), higher flourishing (P = 0.453), higher positive mood (P = 0.518), or lower negative mood (P = 0.538) in the treatment group compared with the control group at follow-up. | CES-D, Mean (SD) | 13.9 (9.9) | 13.6 (10.1) | Effect size: -1.41 (-4.33, 1.50) | 0.339 |
| Dabbaghmanesh, 2019 | The two study groups did not differ significantly in TSH, FT4, and TPO levels at birth (P > 0.05). However, by the fourth postnatal week, depression scores were significantly different between the two groups (P = 0.002). | Edinburgh postnatal depression scale (EPDS) | 8.52 (3.61) | 4.48 (3.30) | NR | <0.0001 |
| Dean, 2011 | Despite significant increases in vitamin D status in the active group, no significant changes were observed in working memory (F = 1.09, p = 0.30), response inhibition (F = 0.82, p = 0.37), cognitive flexibility (F = 1.37, p = 0.24), or secondary outcomes (depression, anxiety, anger). No serious adverse effects were reported. | The Beck Depression Inventory (BDI) | 5.72 (0.83) | 7.24 (0.84) | NR | 0.51 |
| Heidari, 2024 | At the end of the intervention, the mean score of total PMS symptoms showed significant improvement in the vitamin-supplemented group compared with the control group (p < 0.001). When PMS symptoms were divided into five subgroups, the mean score of all subgroups decreased post-supplementation compared with baseline. The greatest decrease was observed in the depression subgroup (53%), while the smallest was in the water retention subgroup (28%). This indicates a greater improvement in mood-related symptoms compared with physical symptoms in this study (p < 0.001). | Premenstrual syndrome (PMS) symptoms assessment | 8.5 (3) | 6 (2.5) | NR | <0.001 |
| Högberg, 2012 | After vitamin D supplementation, well-being increased (p < 0.001), and there was a significant improvement in eight of the nine items on the vitamin D deficiency scale: depressed feeling (p < 0.001), irritability (p < 0.05), tiredness (p < 0.001), mood swings (p < 0.01), sleep difficulties (p < 0.01), weakness (p < 0.01), ability to concentrate (p < 0.05), and pain (p < 0.05). Depression, as measured by the MFQ-S, also showed significant improvement (p < 0.05). | Depressed feelings | 6.5 (2.7) | 4.3 (2.9) | NR | <0.001 |
| Kerr, 2015 | Additionally, the null findings regarding dietary intake of vitamin D are consistent with the notion that even foods fortified or naturally rich in vitamin D make negligible contributions to blood levels compared with season (sunlight exposure) or vitamin supplement use (Holick et al., 2011). Thus, supplementation may be relevant for some populations year-round. | Depressive symptoms | NR | NR | Est. 0.04 (0.09) | β = 0.04; p>0.05 |
| Krysiak, 2018 | Vitamin D supplementation decreased the total BDI-II score in women with vitamin D deficiency. In women with vitamin D insufficiency, supplementation tended to reduce the BDI-II score (p = 0.075) and the percentage of patients with total (p = 0.065) and mild (p = 0.065) depressive symptoms, but it did not affect the percentage of women with moderate or severe depressive symptoms. | BDI-II score, Mean (SD) | Women with vitamin D deficiency beginning of the study<br>13.4 (3.8) | Women with vitamin D after treatment<br>9.1 (3.6) |  | <0.01 |
| Libuda, 2020 | Although the intervention led to a greater increase in 25(OH)D levels in the VG than in the PG (treatment difference: +14 ng/mL; 95% CI, 4.86–23.77; p = 0.003), the change in BDI-II scores did not differ between groups (+1.3; 95% CI, -2.22 to 4.81; p = 0.466). | BDI-II | 29.2 (9.38) | 31.1 (11.3) | 1.29 (-2.22, 4.81) | 0.466 |
| Miyake, 2015 | The prevalence of depressive symptoms during pregnancy was 19.3%. Higher dietary vitamin D intake was significantly associated with a lower prevalence of depressive symptoms during pregnancy, independent of potential dietary and non-dietary confounding factors. | Depressive symptoms | NR | NR | Crude: <0.001<br>Adjusted: 0.02 | p (for trend) = 0.02 |
| Ostadmohammadi, 2019 | Vitamin D and probiotic co-supplementation, compared with placebo, significantly improved Beck Depression Inventory scores [β (difference in the mean outcome between treatment groups) = -0.58; 95% CI, -1.15 to -0.02; p = 0.04], General Health Questionnaire scores (β = -0.93; 95% CI, -1.78 to -0.08; p = 0.03), and Depression, Anxiety, and Stress Scale scores (β = -0.90; 95% CI, -1.67 to -0.13; p = 0.02). | BDI total scores | 13.2 (3.7) | 11.9 (3.4) | -0.58 (-1.15, -0.02) | 0.04 |
| Rajabi-Naeeni, 2021 | There was also a significant difference between the group receiving both supplements and the group receiving only placebos in terms of reductions in depression and stress ($p < 0.05$). | Depression | 13.54 (11.27) | 10.25 (10.16) | NR | Adjusted: $<0.05$ |
| Rouhi, 2018 | Vitamin D supplementation decreased depression and fatigue scores in the intervention group ($p < 0.001$). | Depression | 13.4 (4.7) | 8.6 (4.3) | NR | $<0.001$ |
| Satyanarayana, 2024 | Comparing Beck Depression Inventory scores before and after the intervention showed a statistically significant reduction in the vitamin D intervention group. | BDI-II | Before vitamin D supplementation<br>20.4 (9.2) | After vitamin D supplementation<br>17.0 (7.3) | NR | 0.001 |
| Tsunoda, 2023 | Postpartum depressive symptoms one month after delivery were assessed using the Edinburgh Postnatal Depression Scale. Logistic regression analysis showed a reduced risk of postpartum depressive symptoms for all quintiles of vitamin D intake except the first: second quintile (adjusted odds ratio [95% confidence interval], 0.88 [0.82–0.94]); third (0.83 [0.78–0.89]); fourth (0.87 [0.81–0.93]); and fifth (0.90 [0.83–0.97]). Post-adjustment trend tests revealed a significant association between dietary vitamin D intake and postpartum depressive symptoms ($p$ for trend = 0.004). | Edinburgh postnatal depression scale (EPDS) | NR | NR | Crude odds ratio: Quintile of vitamin D intake; 5 (high) = 0.94 (0.89–1.01)<br>Adjusted odds ratio: Quintile of vitamin D intake; 5 (high) = 0.90 (0.83–0.97) | 0.004 |

For the between-group meta-analysis (n=9), we found that vitamin D supplementation led to a significant reduction in depression scores (e.g., follow-up score - baseline score) compared to placebo (SMD: -0.43, 95% CI: -0.75 to -0.12, p = 0.007, I^2^ = 78%) (**Figure 2A**).^47–53,66,67^ These studies revealed high mean depression scores for baseline compared to follow-up scores for vitamin D supplementation groups, with the exception of two studies.^47,48^ For the within-group meta-analysis (n=10), we found that vitamin D supplementation significantly reduced depression scores within the vitamin D supplementation group compared to baseline (SMD: -1.00, 95% CI: -1.60 to -0.41, p = 0.001, I^2^ = 93%) (**Figure 2B)**.

**Figure 2:**
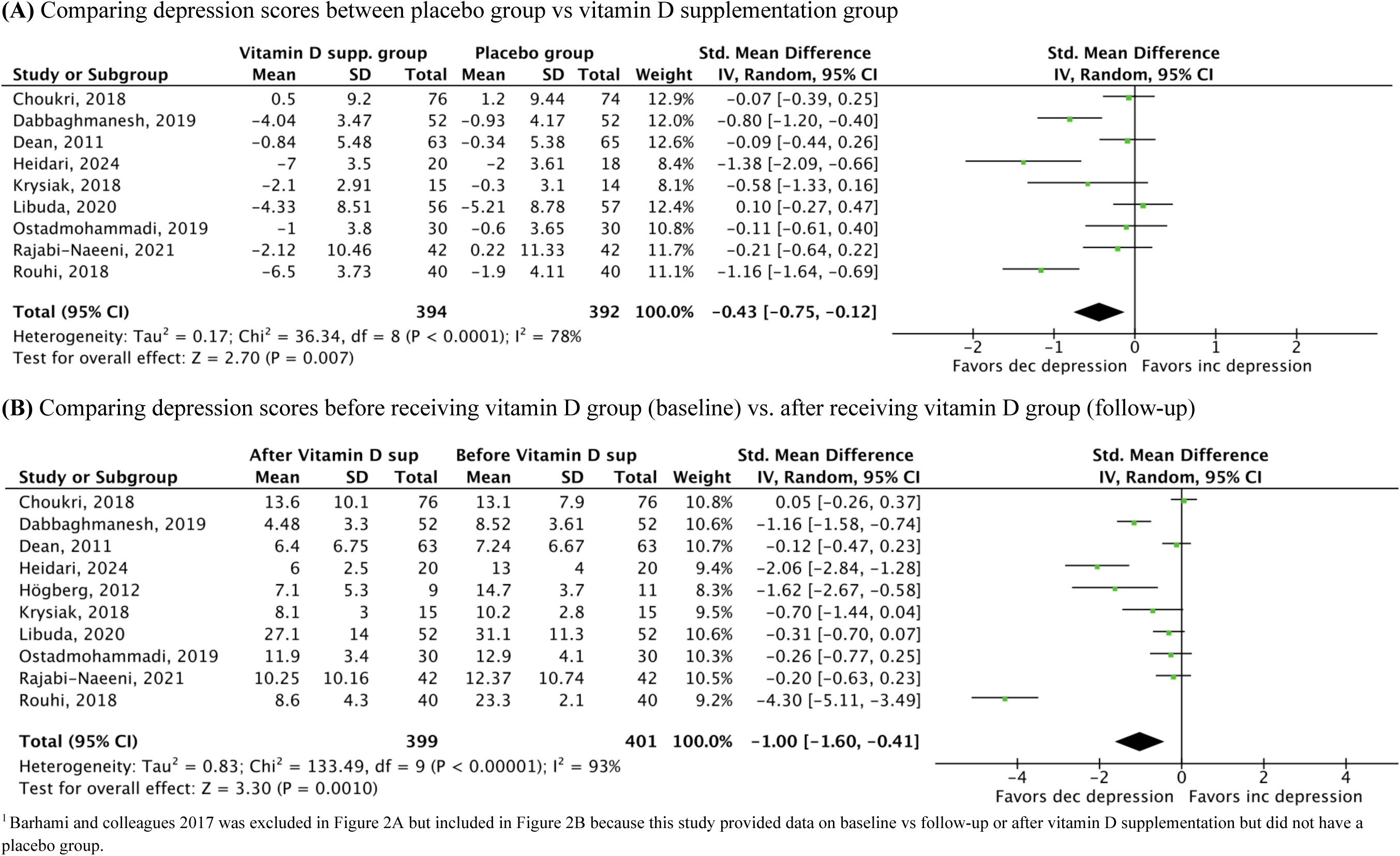
Forest plot of association between vitamin D supplementation and depression among adolescents, young adults, and women during postpartum^1^

### Subgroup analyses

#### Association between vitamin D supplementation and postpartum depression

Six studies specifically investigated the association between vitamin D supplementation and postpartum depression.^47,48,52,53,70,71^ Studies were conducted in New Zealand (n = 1, 17%), Iran (n = 3, 50%), and Japan (n = 2, 33%). Four studies provided sufficient continuous data for meta-analysis. Our meta-analysis revealed that vitamin D supplementation led to a significant decrease in depression scores compared to placebo (SMD: -0.55, 95% CI: -1.04 to -0.06, p = 0.03, I^2^ = 83%) (**Figure 3A)**. When we compared the change in depression score Baseline vs. Follow-up within the vitamin D supplementation group, all studies, with the exception of one study,^47^ found that vitamin D supplementation significantly reduced depression scores within the vitamin D supplementation group compared to baseline (SMD: -1.35, 95% CI: -2.67 to -0.03, p = 0.05, I^2^ = 97%) (**Figure 3B)**.

**Figure 3:**
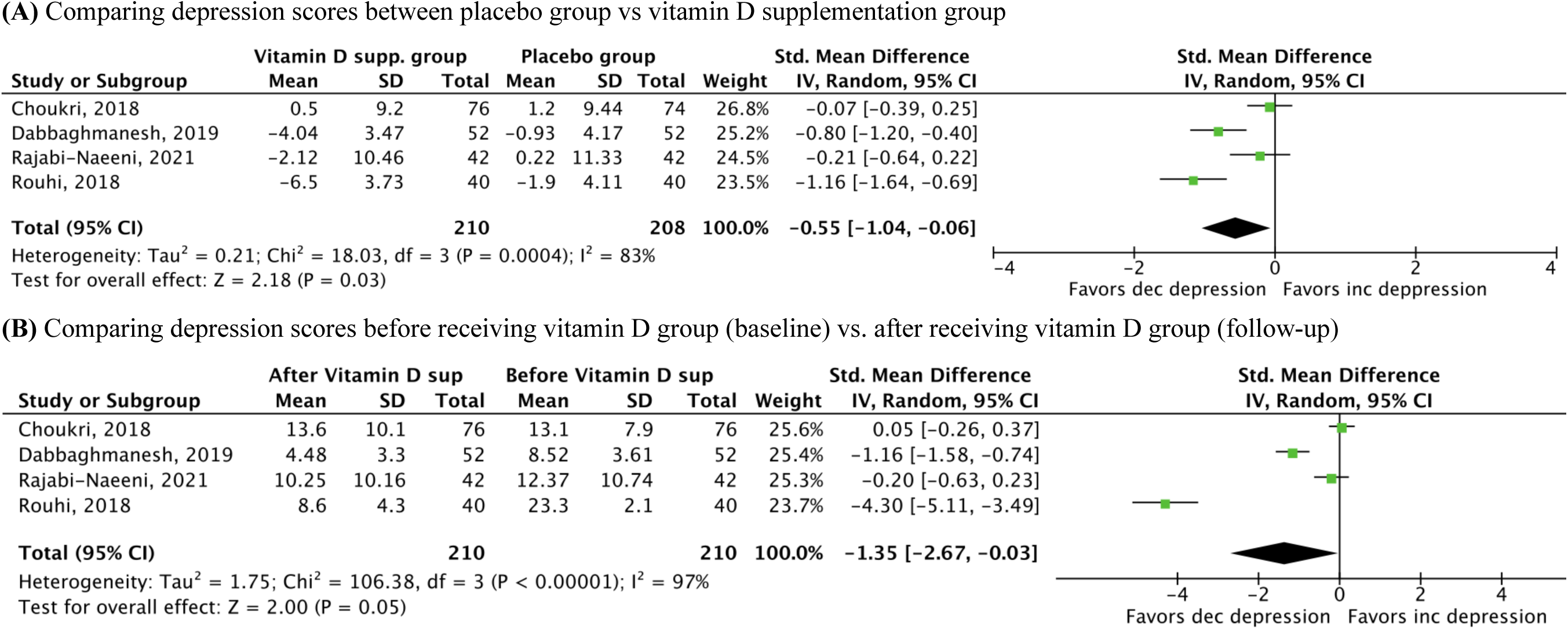
Forest plot of association between vitamin D supplementation and depression among women during postpartum

#### Association between vitamin D and depression among young adults

Our meta-analysis consisted of five studies that investigated the association between vitamin D supplementation and depression among adolescents (n = 1, 20%)^67^ and young adults (n = 4, 80%).^49–51,66^ Studies were conducted in Australia (n = 1, 20%), Iran (n = 2, 40%), Poland (n = 1, 20%), and Germany (n = 1, 20%). Our meta-analysis revealed that vitamin D supplementation led to a significant decrease in depression scores compared to placebo (SMD: -1.38, 95% CI: - 1.65 to -1.10, p < 0.001, I^2^ = 76%) (**Figure 4A)**. When we compared the change in depression score Baseline vs. Follow-up within the vitamin D supplementation group, all studies, with the exception of one study,^49^ found that vitamin D supplementation significantly reduced depression scores within the vitamin D supplementation group compared to baseline (SMD: -0.73, 95% CI: -1.46 to 0.00, p = 0.05, I^2^ = 85%) (**Figure 4B)**.

**Figure 4:**
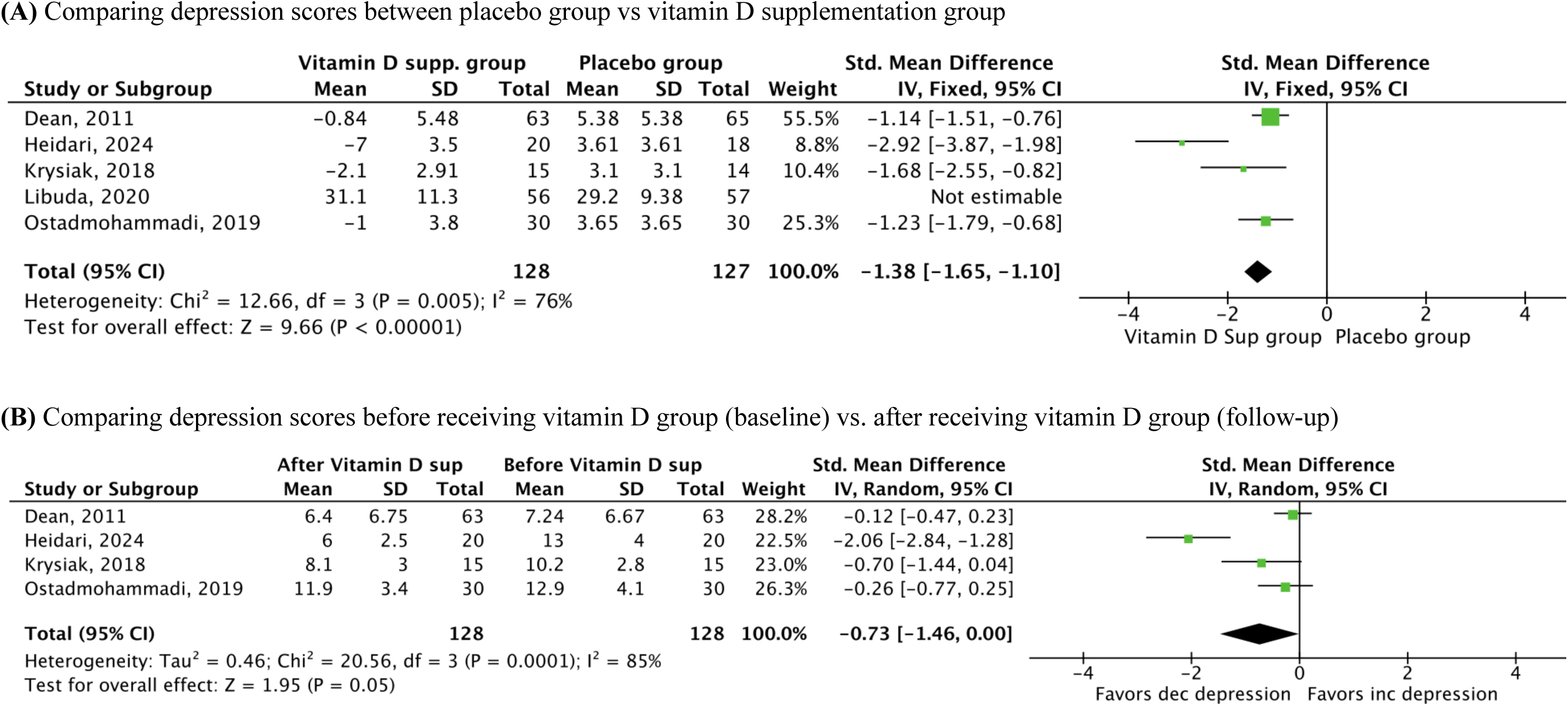
Forest plot of association between vitamin D supplementation and depression among adolescents and young adults

#### Association between vitamin D and depression among adolescents

Four studies investigated the association between vitamin D supplementation and depression among adolescents.^65,67,68,72^ These studies were conducted in Iran (n = 1, 25%), Sweden (n = 1, 25%), Germany (n = 1, 25%), and India (n = 1, 25%). For depression scales, these studies used the Beck Depression Inventory (BDI) (n = 3, 75%) and the Mood and Feelings Questionnaire short version (MFQ-S) (n = 1, 25%). Of the four studies, the majority (n = 3, 75%) found that vitamin D supplementation led to a significant decrease in depression scores.^65,68,72^ These studies did not provide sufficient continuous data to conduct a meta-analysis.

### Secondary outcome

#### Anxiety measurement instruments

Of the fifteen included studies, we found four studies that used “anxiety” as a secondary outcome and provided sufficient data to conduct a meta-analysis.^47,49,52,66^ These studies used the Hospital Anxiety and Depression Scale (n = 1, 25%),^47^ the State-Trait Anxiety Inventory (STAI) (n = 1, 25%),^49^ the Beck’s Anxiety Inventory (BAI) (n = 1, 25%),^66^ and the Depression Anxiety Stress Scale-21 (DASS-21) (n = 1, 25%) (see **Appendix 5** for list of validated anxiety measures).^52^ These studies were conducted in New Zealand (n = 1, 25%), Australia (n = 1, 25%), and Iran (n = 2, 50%).

#### Association between vitamin D supplementation and anxiety

Four studies provided sufficient continuous data for meta-analysis. We found that vitamin D supplementation led to a no significant reduction in anxiety scores compared to placebo (SMD: - 0.31, 95% CI: -0.75 to 0.12, p=0.16, I^2^ = 77%) (**Figure 5A**). When we compared the change in anxiety scores within the vitamin D supplementation group (e.g., baseline anxiety scores vs. follow-up anxiety scores) across studies, we found that vitamin D supplementation did not significantly reduce anxiety scores compared to baseline (SMD: -0.41, 95% CI: -1.03 to 0.21, p=0.19, I^2^ = 88%) (**Figure 5B)**.

**Figure 5:**
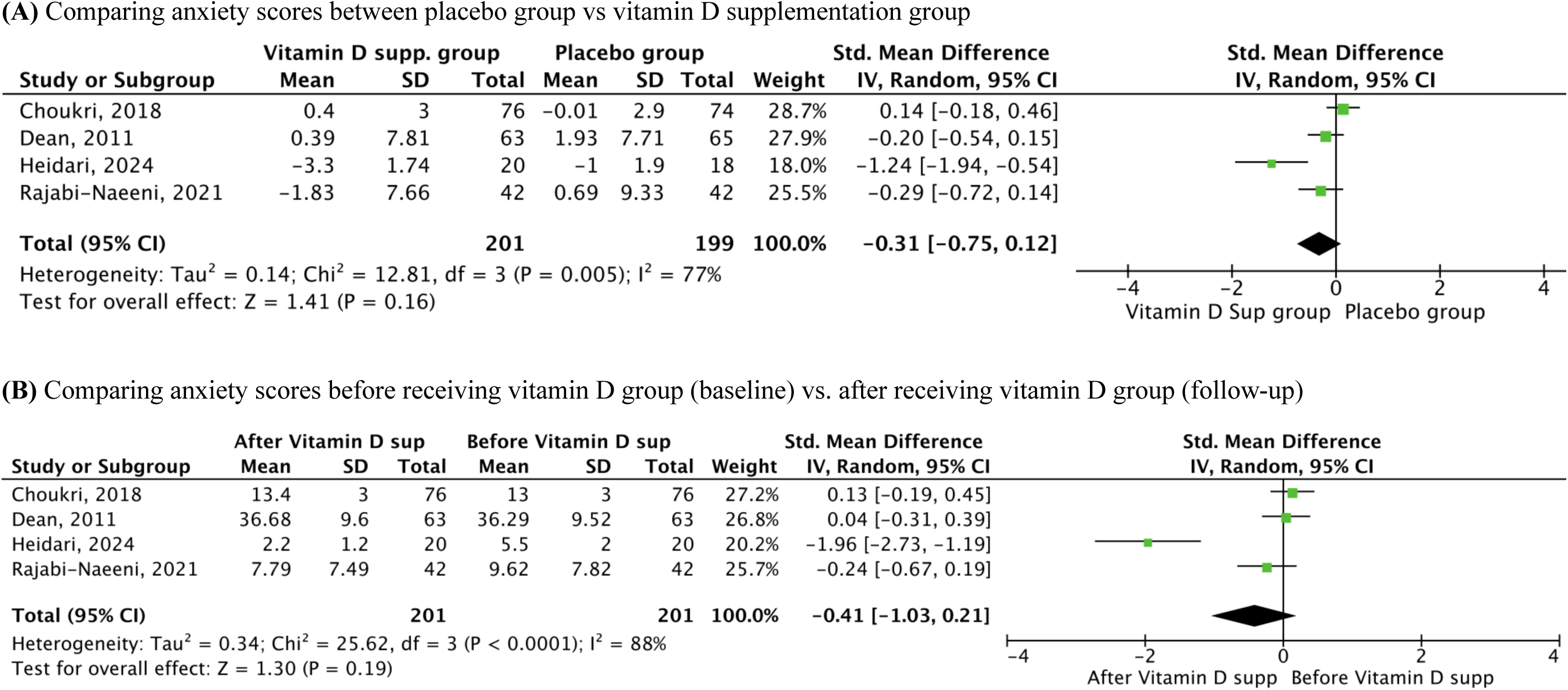
Forest plot of association between vitamin D supplementation and anxiety among young adults, including women during postpartum

### Publication bias

Our funnel plot of the primary outcome reveals a symmetrical distribution of studies, which indicates little to no publication bias in the included studies (**Figure 6**).

**Figure 6.**
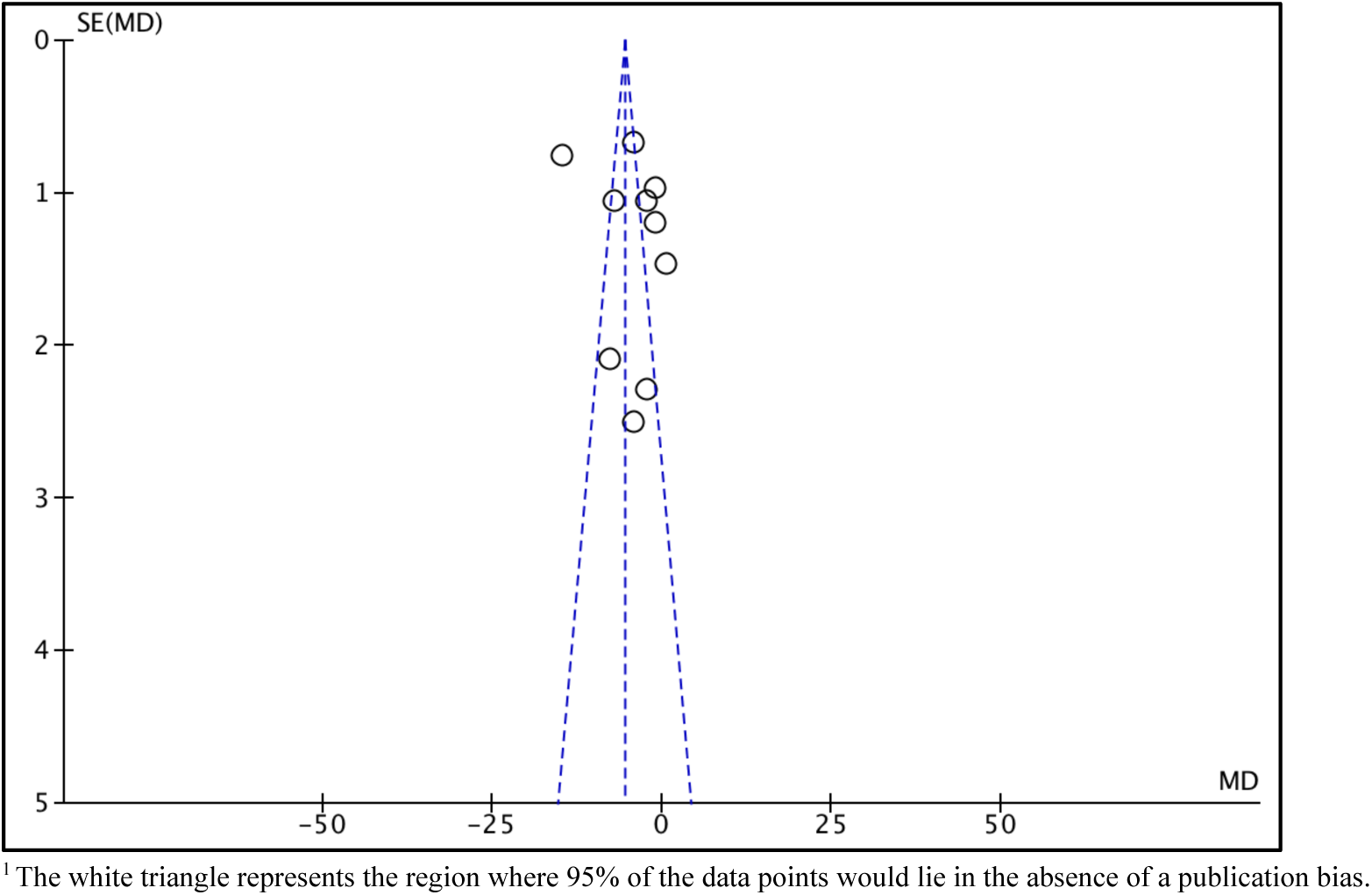
Funnel plot of meta-analysis of published studies that reported the impact of vitamin D supplementation on depression scores before (baseline) vs. after receiving vitamin D supplementation (follow-up).^1^

## DISCUSSION

### Key findings

Based on the 15 studies included in this systematic review, the majority reported an association between vitamin D supplementation and a reduction in depression scores among adolescents and young adults. Our meta-analysis revealed four significant results: (1) vitamin D supplementation led to a significant decrease in depression scores during follow-up compared with baseline and with placebo in the overall population of interest; (2) in subgroup analyses, a significant association was found both compared with baseline and with placebo among women during the postpartum period; (3) among young adults, a significant association was observed only when compared with placebo; and (4) vitamin D supplementation was not associated with a decrease in anxiety scores when compared with either placebo or baseline.

### Results in context

Overall, the findings of our review align with previous evidence and contribute to the ongoing discussion on the potential role of vitamin D supplementation in alleviating depressive symptoms.^73–75^ Our review incorporated several recently published studies that were not included in prior systematic reviews on this topic, and the majority of these studies reported a significant benefit of vitamin D supplementation in reducing depressive symptoms, particularly among adolescents and young adults.^48,52,66–68,71^ Moreover, population-based studies have suggested that vitamin D supplementation may be particularly effective in individuals with moderate to severe depressive symptoms or those with underlying deficiencies. However, while the majority of the studies (n = 11) included in our review supported a beneficial effect of vitamin D supplementation on depression, the remaining studies (n = 4) found no significant association, consistent with earlier trials that reported mixed findings on this relationship.^76^ These inconsistencies may be attributable to differences in study design, dosage, baseline vitamin D levels, comorbidities, and participant characteristics. Therefore, while our results do not fully resolve the debate, they provide valuable insights into the nuanced effects of vitamin D supplementation on depression, emphasizing the importance of considering individual differences such as baseline vitamin D status and severity of depressive symptoms when evaluating potential benefits. Our findings also highlight the need for further well-designed randomized controlled trials to clarify the efficacy of vitamin D supplementation as an adjunctive intervention for depression.

### Quality of the evidence

The methodological quality of the included studies was classified as “good” for all publications. All studies adhered to standardized vitamin D supplementation protocols and used validated diagnostic tools to assess depressive symptoms. However, variability in study design, dosage, duration of supplementation, and participant characteristics may have influenced the findings. Some studies had relatively small sample sizes (e.g., n < 100), which could affect the reliability of the results by increasing the risk of Type I (false positive) and Type II (false negative) errors, ultimately limiting the generalizability of the findings. Larger sample sizes typically enhance statistical power and improve the reliability of conclusions regarding the effects of vitamin D supplementation on depression. Additionally, the limited number of studies (n = 10) available for meta-analysis may constrain the overall strength and precision of the pooled estimates, as larger meta-analyses generally provide more robust and reliable conclusions. These factors underscore the need for future well-powered, high-quality, randomized controlled trials to clarify the potential role of vitamin D supplementation in depression management.

### Strengths and limitations

Our review includes recently published research studies examining the effects of vitamin D supplementation on depression, providing an updated synthesis of the evidence. We incorporated studies conducted both in the United States and internationally, enhancing the generalizability of our findings. A key strength of this review is the rigorous assessment of methodological quality, with all included studies rated as “good” based on the Newcastle-Ottawa Scale, ensuring reliability and rigor in the conclusions drawn. However, one limitation is that studies conducted outside of the United States may introduce cultural and healthcare system differences that could influence responses to vitamin D supplementation and the reporting of depressive symptoms, potentially affecting the applicability of findings to the U.S. population. Additionally, the studies included in our review varied in sample size, ranging from small to large-scale populations, which may impact the consistency of the findings. Another important limitation is that our review does not establish the causal direction of the relationship between vitamin D supplementation and depression. Specifically, while we analyzed the impact of supplementation on depressive symptoms, it remains unclear whether improvements in depression lead to better vitamin D metabolism or whether other confounding factors contribute to the observed effects. Future studies with rigorous randomized controlled trial designs are needed to better elucidate the causal pathways and optimize vitamin D supplementation strategies for depression management.

### Impact, implications, and next steps

The findings of this systematic review suggest that vitamin D supplementation may have a beneficial role in reducing depressive symptoms, particularly among individuals with vitamin D deficiency. These results have potential implications for a broad range of stakeholders, including individuals experiencing depression, healthcare providers, policymakers, pharmaceutical companies, and researchers. Given the observed associations, there may be an opportunity to incorporate routine screening of vitamin D levels in depression assessments, which could influence current clinical practices and healthcare guidelines. If vitamin D supplementation proves to be an effective adjunctive treatment, it could reshape standard approaches to depression management, encouraging healthcare professionals to consider vitamin D status as a modifiable risk factor. Additionally, these findings highlight the need for targeted interventions, particularly in populations at higher risk for both vitamin D deficiency and depression, such as older adults, individuals with limited sun exposure, and those with dietary insufficiencies. Future research should focus on well-designed randomized controlled trials to establish the efficacy of vitamin D supplementation in different populations and to determine optimal dosing regimens. Further studies could also explore how factors such as age, gender, genetic predisposition, and baseline vitamin D status influence the effectiveness of supplementation. Expanding this research may provide more precise guidelines for integrating vitamin D into mental health care strategies and public health policies aimed at improving mental well-being.

## CONCLUSION

In summary, most of the included studies suggest that vitamin D supplementation may have a beneficial effect on depressive symptoms, particularly in individuals with vitamin D deficiency. Our findings indicate potential variations in the effectiveness of supplementation based on factors such as age and gender, highlighting the need for further investigation into these demographic differences. Future research should focus on randomized controlled trials to determine whether vitamin D supplementation consistently demonstrates similar associations with reductions in depression. Additionally, exploring optimal dosing strategies, treatment duration, and individual characteristics that may modify the response to supplementation will be essential in refining clinical recommendations and public health strategies for depression management.

## Review Protocol

We created a protocol in July 2024 to guide the inclusion of studies. We registered our protocol in August 2024 (see **Appendix 7** for systematic review protocol modifications).

## CRediT authorship contribution statement

**Joseph P. Nano:** Conceptualization, methodology, formal analysis, writing – original draft. **William Catterall:** Conceptualization, methodology, writing – original draft**. Salar Khaleghzadegan:** methodology, writing – review and editing. **Camilo A. Castelblanco**: methodology, writing – review and editing. **Heather B. Blunt:** methodology, writing – review and editing. **Renata W. Yen:** Methodology, formal analysis, writing – original draft.

## Data Availability

All data produced in the present work are contained in the manuscript

## Acknowledgment

We would like to thank the faculty and colleagues at The Dartmouth Institute for Health Policy and Clinical Practice, particularly those from the PH100 Inferential Methods in Epidemiology course, for inspiring this work through class discussions.

## APPENDICES

**Appendix 1:** Inclusion criteria table & outcomes of interest.

| Category | Inclusion Criteria | Justification |
| --- | --- | --- |
| Study design | Cohort study | Cohort studies can investigate the association between multiple outcomes and exposures in a single study for an extended period of time. This is important to show changes in vitamin D serum concentration and depression scores over time. |
|  | Case study | Case studies focus on a single individual or group and allow researchers to collect information on unusual cases potentially related to the association between vitamin D and depression. |
|  | Cross-sectional | Cross-sectional studies provide a comparison between different variables, such as sleep, age, LDL levels, vitamin D supplementation, and depression scores at the same time. |
|  | Cross-sectional survey study | Survey study designs are observational, as there is no experimental component. Also, they are useful in recruiting a large number of participants and describing the characteristics of a large population, such as age, race, hours of sleep, and past medical history. |
|  | Pilot study | Pilot studies can be used to provide information about the association between vitamin D and depression by conducting a smaller-scale study to assess the safety of treatment or interventions.<sup>77</sup> |
|  | Randomized control trial study | Randomized control trials (RCT) can be used to investigate whether there is a causal relationship between vitamin D and depression while reducing chances of selection bias.<sup>78</sup> |
| Population | Adolescents<sup>79-81</sup> (defined by $\geq 10$ and $17 \leq$) and young adults<sup>38,39</sup> ($\geq 18$ and $\leq 40$; all pregnant women with reported postpartum depression) | This is the most relevant population to our research topic because depression is more likely to start during adolescence, and this population is less likely to experience morbidity that would increase the risk for depression.<sup>82</sup> If feasible, we would plan to evaluate studies of young adults ($\geq 18$ and $\leq 39$), and perhaps a separately subgroup-analysis of adolescents ($\geq 11$ and $17 \leq$) to extend our findings. The age cutoff for adolescents was based on the World Health Organization's age standard.<sup>81</sup><br>According to the World Health Organization, the age of reproductive women ranges from 15 to 49.<sup>83</sup> When a woman is younger than 30, she has an 85% chance to conceive within one year. This chance declines to 66% at the age of 35 and 44% at the age of 40.<sup>84</sup> Also, 27% of pregnancies end in a miscarriage at age 40 compared to 16% at age 30 or younger).<sup>85</sup> Therefore, for postpartum depression, we assume that the majority of mothers are within the age of range of 18 - 40. |
| Intervention / Exposure | Serum Vitamin D supplementation | According to the Mayo Clinic website, the recommended daily amount of vitamin D supplementation is the following.<sup>86</sup><br>(1) 400 international units (IU) for children up to age 12 months<br>(2) 600 IU for people ages 1 to 70 years<br>(3) 800 IU for people over 70 years.<br>We included various common vitamin D intake administration routes: oral, parenteral, intravenous, and intramuscular vitamin D administration.<sup>87</sup> Also, we included studies that used the intradermal route (direct sunlight |
|  |  | exposure) as long as it met the minimum number of minutes of exposure that matches with the recommended dosage of vitamin D supplementation (approximately 15 minutes of exposure = 1000 IU vitamin D).<sup>88</sup> |
| Comparison condition | Placebo | We included studies with a placebo control group. Or studies that did not use vitamin D supplementation in their comparison group. |
| Primary outcome | Depression | Several depression questionnaires are used to measure depression (**Appendix 5**). |
| Secondary outcome | Anxiety | Several anxiety questionnaires are used to measure anxiety (**Appendix 5**). |

**Appendix 2:** PubMed search terms table.

| Theme | MeSH terms (exploded) | Key words |
| --- | --- | --- |
| Depression | "Depression"[Mesh] OR<br>"Depressive Disorder"[Mesh] | Depression [TIAB] OR Depressive Disorder [TIAB]<br>OR seasonal affective[TIAB] OR Dysthym*[TIAB] OR<br>affective disorder*[TIAB] OR major depress*[TIAB]<br>OR unipolar depress*[TIAB] |
| Young Adults and<br>Adolescent | "Young Adult"[Mesh] OR<br>"Adolescent"[Mesh] | Young Adult* [TIAB] OR Adolescen* [TIAB] OR<br>Teen* [TIAB] |
| Vitamin D | "Vitamin D"[Mesh] OR "Vitamin<br>D Deficiency"[Mesh] OR<br>Hydroxycholecalciferols[Mesh]<br>OR cholecalciferol[Mesh] OR<br>ergocalciferol[Mesh] | Vitamin D* [TIAB] OR hydroxycholecalciferol*[TIAB]<br>OR 25-hydroxyvitamin D[TIAB] OR<br>cholecalciferol*[TIAB] OR ergocalciferol*[TIAB] OR<br>calcifediol*[TIAB] OR calcitriol*[TIAB] OR<br>hydroxyvitamin*[TIAB] |

**Appendix 3:** Search strategies^1^.

| Database | Date | Search | Query | Results |
| --- | --- | --- | --- | --- |
| MEDLINE via PubMed | 1946 to May 29, 2024 | #4 | Search: #1 AND #2 AND (english[Filter]) | 1,418 |
|  |  | #3 | Search: #1 AND #2 | 1,470 |
|  |  | #2 | Search: "Vitamin D"[Mesh] OR "Vitamin D Deficiency"[Mesh] OR Hydroxycholecalciferols[Mesh] OR cholecalciferol [Mesh] OR ergocalciferol [Mesh] OR Vitamin D [TIAB] OR Vitamin D1 [TIAB] OR Vitamin D2 [TIAB] OR Vitamin D3 [TIAB] OR hypovitaminosis d [TIAB] OR hydroxycholecalciferol*[TIAB] OR 25-hydroxyvitamin D[TIAB] OR cholecalciferol*[TIAB] OR ergocalciferol*[TIAB] OR calcifediol*[TIAB] OR calcitriol*[TIAB] OR hydroxyvitamin*[TIAB] | 117,550 |
|  |  | #1 | Search: "Depression"[Mesh] OR "Depressive Disorder"[Mesh] OR Depression [TIAB] OR Depressive Disorder [TIAB] OR seasonal affective[TIAB] OR Dysthym*[TIAB] OR affective disorder*[TIAB] OR major depress*[TIAB] OR unipolar depress*[TIAB] | 540,624 |
| The Cochrane Library | 1898 to May 29, 2024 | #3 | #1 AND #2 | 620 |
|  |  | #2 | ("Vitamin D" OR "Vitamin D deficiency" OR "Vitamin D1" OR "Vitamin D2" OR "Vitamin D3" OR hydroxycholecalciferol* OR "25-hydroxyvitamin D" OR cholecalciferol* OR ergocalciferol* OR calcifediol* OR calcitriol* OR hydroxyvitamin* OR "hypovitaminosis d"):ti,ab,kw (Word variations have been searched) | 17,756 |
|  |  | #1 | (Depression OR "Depressive Disorder" OR "seasonal affective" OR Dysthym* OR (Affective NEXT disorder*) OR (major NEXT depress*) OR (unipolar NEXT depress*)):ti,ab,kw (Word variations have been searched) | 117,803 |
| CINAHL | 1981 to May 29, 2024 | #4 | S1 AND S2 AND narrow by Language: English | 805 |
|  |  | #3 | S1 AND S2 | 824 |
|  |  | #2 | ((MH "Vitamin D Deficiency+") OR (MH "Ergocalciferols") OR (MH "Vitamin D+") OR (MH "Cholecalciferol") ) OR ( "Vitamin D" OR "Vitamin D1" OR "Vitamin D2" OR "Vitamin D3" OR "hypovitaminosis d" OR hydroxycholecalciferol* OR "25- | 32,432 |
|  |  |  | hydroxyvitamin D" OR cholecalciferol* OR ergocalciferol* OR calcifediol* OR calcitriol* OR hydroxyvitamin* ) |  |
|  |  | #1 | (MH "Depression+") OR ( Depression OR "Depressive Disorder" OR "seasonal affective" OR Dysthym* OR "affective disorder*" OR "major depress*" OR "unipolar depress*" ) | 223,268 |
| PsychINFO | 1806 to May 29, 2024 | #4 | S1 AND S2 AND narrow by Language: English | 486 |
|  |  | #3 | S1 AND S2 | 499 |
|  |  | #2 | "vitamin d" OR "Vitamin D1" OR "Vitamin D2" OR "Vitamin D3" OR "hypovitaminosis d" OR "hydroxycholecalciferol*" OR "25-hydroxyvitamin D" OR cholecalciferol* OR ergocalciferol* OR calcifediol* OR calcitriol* OR hydroxyvitamin* | 2,605 |
|  |  | #1 | ( (DE "Depression (Emotion)") AND (DE "Major Depression" OR DE "Anaclitic Depression" OR DE "Dysthymic Disorder" OR DE "Endogenous Depression" OR DE "Late Life Depression" OR DE "Postpartum Depression" OR DE "Reactive Depression" OR DE "Recurrent Depression" OR DE "Treatment Resistant Depression" OR DE "Atypical Depression" OR DE "Persistent Depressive Disorder") ) OR ( Depression OR "Depressive Disorder" OR "seasonal affective" OR Dysthym* OR "affective disorder*" OR "major depress*" OR "unipolar depress*" ) | 445,667 |
| ClinicalTrials.gov | 1997 to May 29, 2024 | #1 | We searched ClinicalTrials.gov using the term "Depression" as a condition and the terms "Vitamin D" | 56 |
| Scopus | 1970 to May 29, 2024 | #1 | ( TITLE-ABS-KEY ( depression OR "Depressive Disorder" OR "seasonal affective" OR dysthym* OR "major depress*" OR "unipolar depress*" ) ) AND ( TITLE-ABS-KEY ( "Vitamin D" OR "Vitamin D1" OR "Vitamin D2" OR "Vitamin D3" OR "hypovitaminosis d" OR hydroxycholecalciferol* OR "25-hydroxyvitamin D" OR cholecalciferol* OR ergocalciferol* OR calcifediol* OR calcitriol* OR hydroxyvitamin* ) ) AND ( LIMIT-TO ( LANGUAGE , "English" ) ) | 4,253 |

<sup>1</sup> All searches were limited to English

**Appendix 4:**
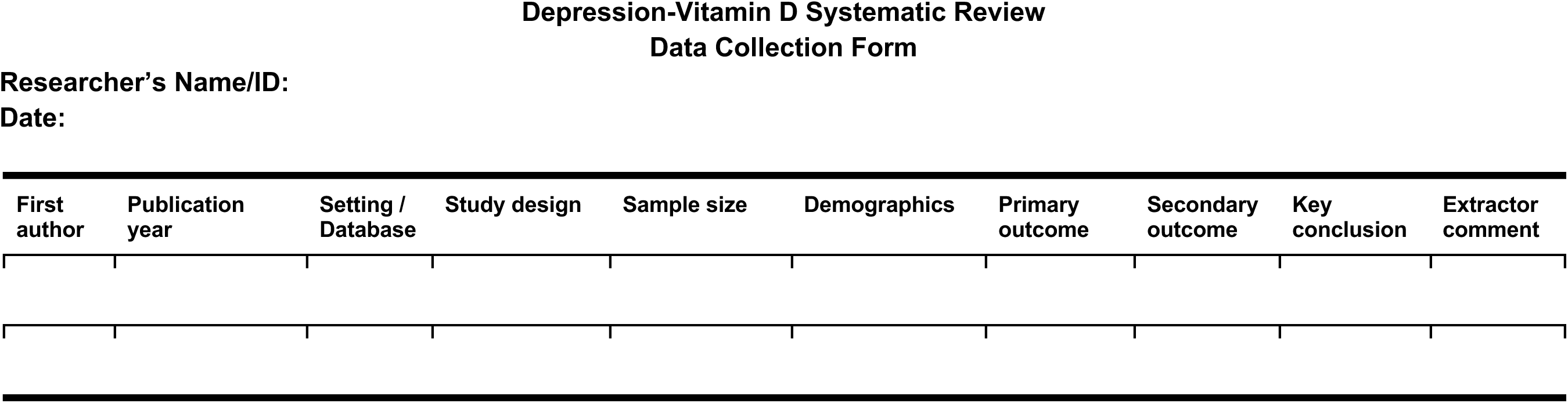
Data collection form.

**Appendix 5:**
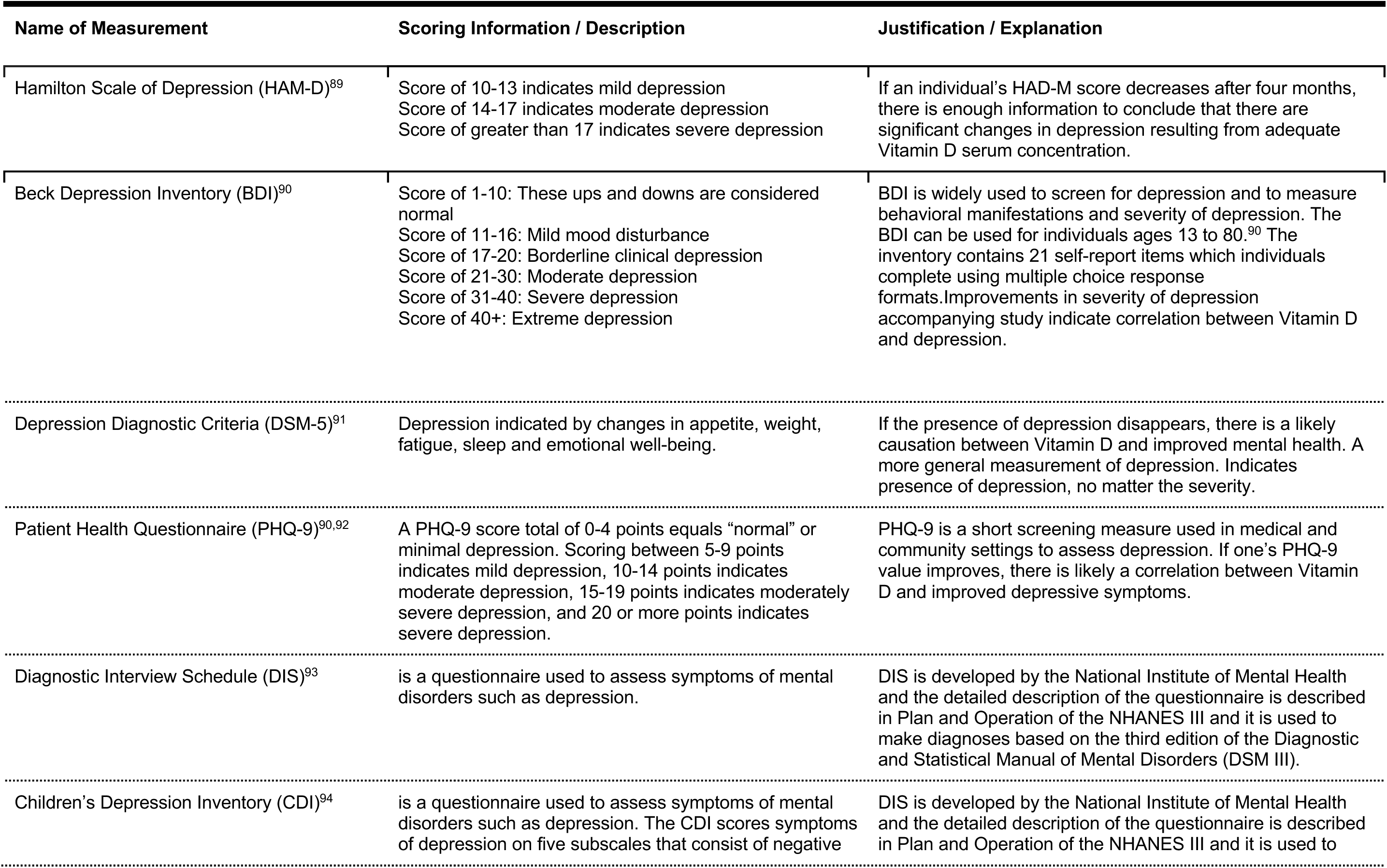
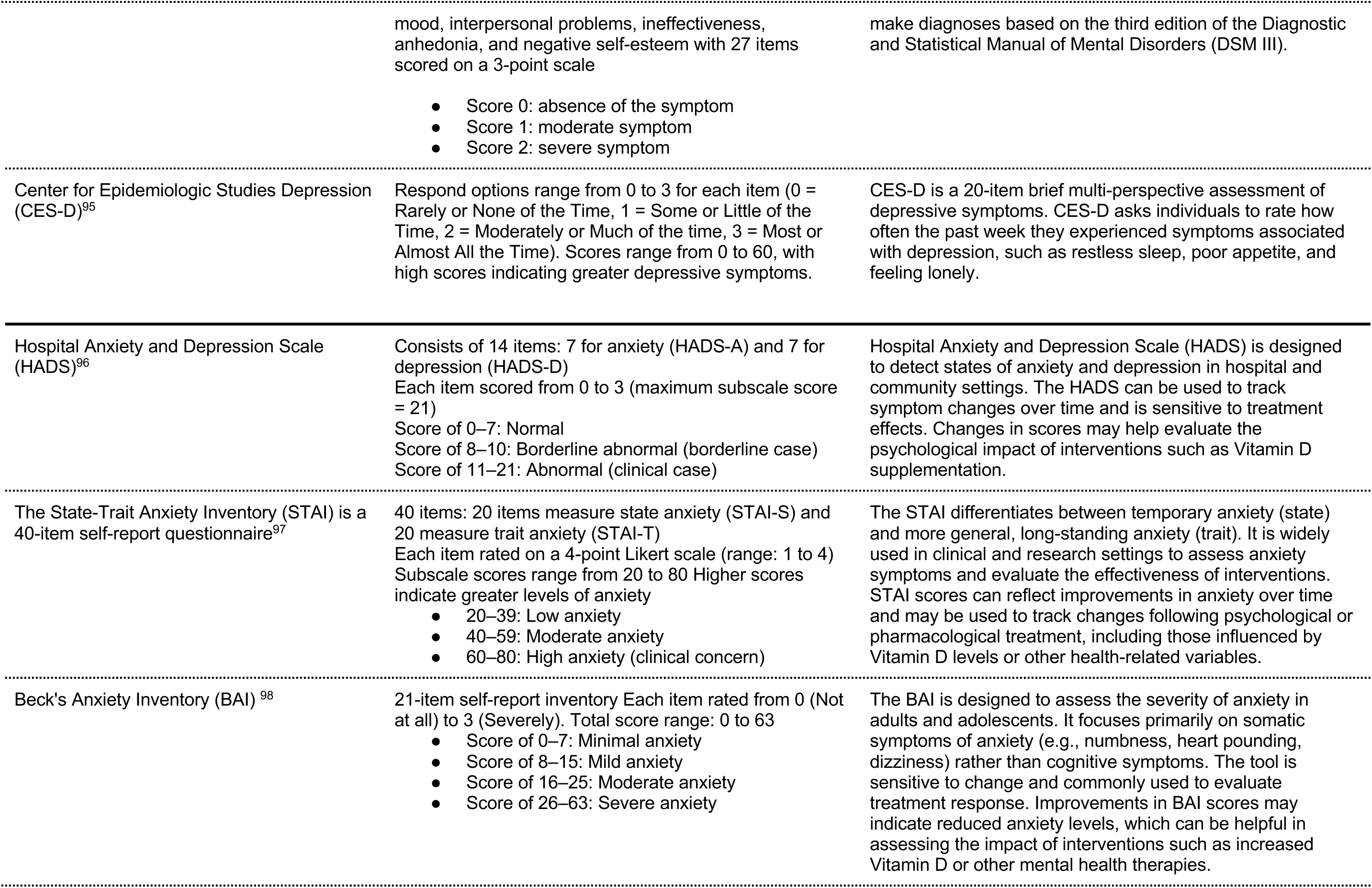
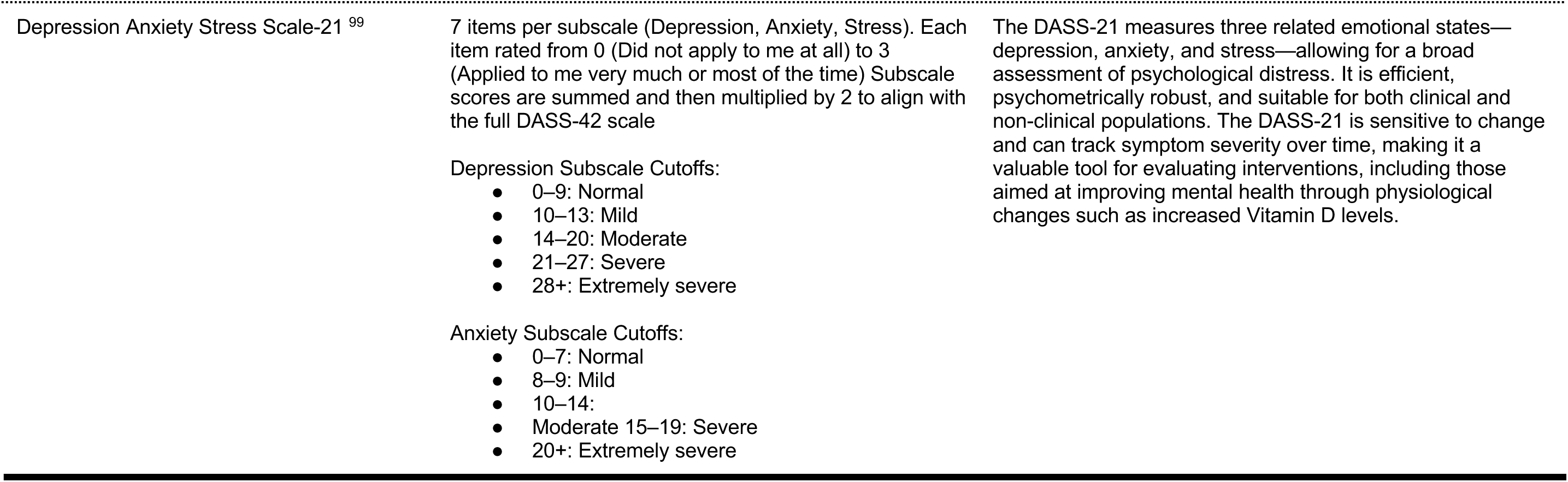
Included validated depression and anxiety screening tools.

**Appendix 6:**
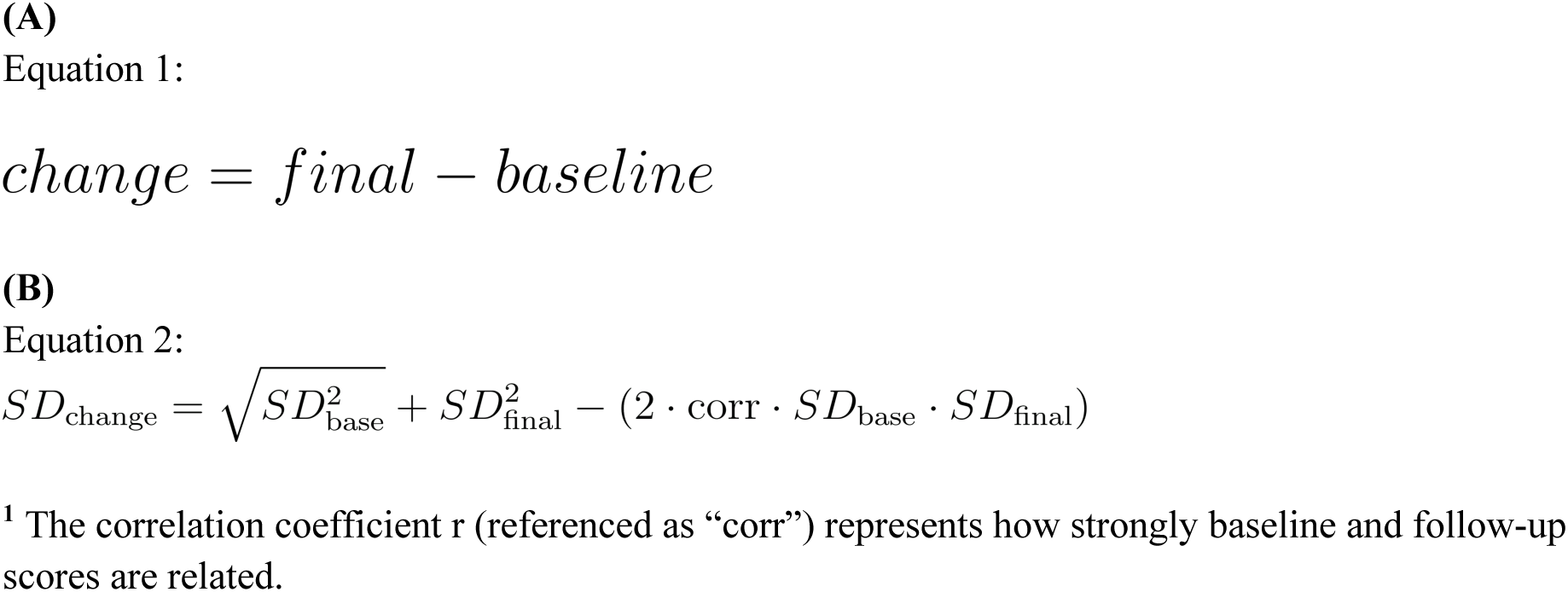
Mathematical equations used to compute (A) mean change from baseline and (B) standard deviations ^1^.

**Appendix 7:** Systematic Review Protocol Modifications.

| Date | Modification | Justification |
| --- | --- | --- |
| July 31, 2024 | Limited to studies in English | To avoid any bias that can happen during translation |

